# Analysis of the potential for a malaria vaccine to reduce gaps in malaria intervention coverage

**DOI:** 10.1101/2020.10.09.20209973

**Authors:** H. Juliette T. Unwin, Lazaro Mwandigha, Peter Winskill, Azra C. Ghani, Alexandra B. Hogan

## Abstract

**Background:** The RTS,S/AS01 malaria vaccine is currently being piloted in three African countries. We sought to identify whether vaccination could reach additional children who are at risk from malaria but do not currently have access to, or use, core malaria interventions.

**Methods:** Using data from household surveys we calculated the overlap between malaria intervention coverage and childhood vaccination (diphtheria-tetanus-pertussis dose 3, DTP3) uptake in 20 African countries with at least one first administrative level unit with *Plasmodium falciparum* parasite prevalence greater than 10%. We used multilevel logistic regression to explore patterns of overlap by demographic and socioeconomic variables. We also estimated the public health impact of delivering RTS,S/AS01 to those children who do not use an insecticide-treated net (ITN) but who received the DTP3 vaccine.

**Results:** Uptake of DTP3 was higher than malaria intervention coverage in most countries. Overall, 34% of children did not use ITNs and received DTP3, while 35% of children used ITNs and received DTP3, although this breakdown varied by country. We estimated that there are 33 million children in these 20 countries who do not use an ITN. Of these, 23 million (70%) received the DTP3 vaccine. Vaccinating those 23 million children who receive DTP3 but do not use an ITN could avert an estimated 9.7 million clinical malaria cases each year. An additional 10.8 million cases could be averted by vaccinating those 24 million children who receive the vaccine and use an ITN. Children who had access to or used an ITN were 9 to 13% more likely to reside in rural areas compared to those who had neither intervention regardless of vaccination status. Mothers’ education status was a strong predictor of intervention uptake and was positively associated with use of ITNs and vaccination uptake and negatively associated with having access to an ITN but not using it. Wealth was also a strong predictor of intervention coverage.

**Conclusions:** Childhood vaccination to prevent malaria has the potential to reduce inequity in access to existing malaria interventions and could substantially reduce the childhood malaria burden in sub-Saharan Africa, even in regions with lower existing DTP3 coverage.

## Background

The introduction of the Millennium Development Goals in 2000 helped to catalyse widespread scale-up of core contemporary malaria control interventions in sub-Saharan Africa: insecticide treated nets (ITNs), indoor residual spraying (IRS), chemoprevention for pregnant women, and more recently, chemoprevention of children in areas of seasonal transmission [1]. Access to treatment of clinical malaria with artemisinin-based combination therapy also increased [1]. Although malaria burden has declined significantly over the previous two decades, malaria remains a leading cause of childhood morbidity and mortality. Worldwide in 2018 there were an estimated 228 million cases of malaria and 405,000 deaths, with over 90% of the deaths occurring in sub-Saharan Africa [2]. Funding for malaria has remained relatively stable since 2010, but the level of investment remains far short of what is required under the WHO Global Technical Strategy for Malaria (GTS) [3]. The first milestone for burden and mortality reduction under the GTS, namely a reduction of at least 40% in malaria incidence and mortality globally by 2020 compared with 2015 levels [3], is therefore unlikely to be met.

Between 2016 and 2018, 578 million ITNs were delivered globally, compared to a total of 582 million between 2014 and 2016 [2, 4]. However, reported usage of ITNs has improved only marginally since 2015 [2]. In addition, fewer people at risk of malaria are being protected by IRS; globally, IRS protection declined from a peak of 5% in 2010 to 2% in 2018 [2]. Furthermore, a high proportion of febrile children do not receive medical care (median: 36%, IQR: 28–45% based on a separate analysis of 20 household surveys conducted in sub-Saharan Africa between 2015 and 2018) [2]. Uptake of these interventions varies within and between countries and is not evenly distributed across demographic and socioeconomic strata [5–7]. While vector control has generally been identified as being more equitable – with ITN distribution in particular identified as “pro-poor” – wealth inequities persist [5, 8] and Webster et al. [7] found that ever-treated net coverage was strongly biased towards richer households in almost all countries included in their study.

Vaccination is one of the most successful and cost-effective public health measures, and has the potential to greatly reduce inequity, particularly in low- and middle-income countries [9, 10]. The Expanded Programme on Immunization (EPI) has been a key catalyst in expanding access to childhood vaccines, with an estimated 86% coverage of DTP3 globally and 78% coverage in Africa in 2018 [11]. However, gaps in coverage do remain, with higher levels of inequality in countries with lower vaccine uptake and evidence of “pro-rich” coverage in some countries [12, 13]. Identifying those that are not receiving basic vaccines is therefore a priority [14, 15]. The majority of children who remain unvaccinated are geographically concentrated, with 60% of these children residing in ten countries (including four malaria endemic countries in Africa: Angola, the Democratic Republic of Congo, Ethiopia, and Nigeria) [14].

The RTS,S/AS01 vaccine for *P. falciparum* malaria is the first vaccine to show partial protection against clinical and severe malaria in children. The multi-site phase 3 trial of the RTS,S vaccine demonstrated 39.0% (95% CI 34.3–43.3%) protective efficacy against clinical malaria in young children who received all four doses, over four years of follow-up according to the per-protocol population [16]. Pilot implementation of the vaccine is now underway in three African countries – Ghana, Kenya and Malawi [17] – and the findings from this pilot will inform public health policy decisions about wider roll-out of the vaccine, including the potential for RTS,S to be incorporated in the EPI [18].

In this study we sought to quantify the overlap and gaps between malaria intervention coverage and childhood vaccination uptake in malaria-endemic African countries. Using data from household-based Demographic and Health Surveys (DHS), we stratified individual children into groups on the basis of reported ITN coverage (ownership and usage) and DTP3 vaccination status [19]. We used these groupings to explore socio-economic factors driving intervention uptake and to quantify the potential that introduction of the RTS,S vaccine could have in reducing malaria burden in those who are not currently accessing or using ITNs.

## Methods

### Data sources

Data were obtained from DHS and Malaria Indicator Surveys (MIS) in Africa [19]. Both are large, nationally representative, household surveys typically conducted every three to four years. In our initial scope we included all African countries with at least one administrative 1 (admin-1) unit with *P. falciparum* parasite prevalence in 2–10-year-old individuals (*Pf*PR_2-10_) greater than 10% based on Malaria Atlas Project (MAP) estimates for 2016 [20]. This threshold was chosen as the level above which the RTS,S malaria vaccine has been estimated to be highly cost-effective [21]. Twenty-six countries fitted our inclusion criteria. For each, we identified the most recent DHS and MIS where geolocation data and parasite prevalence data were also available. Geolocation data were not available for South Sudan, Equatorial Guinea or Niger, and prevalence data were not available for Chad, Gabon or the Central African Republic. These countries were therefore excluded from the analysis. This resulted in 20 countries eligible for analysis with a median country-level parasite prevalence of *Pf*PR_2-10_ = 23% (range 5–43%). The DHS and/or MIS dataset used for each country is listed in the (Table S1).

To quantify vaccination uptake, we extracted individual-level data on the childhood vaccination status of DTP3 (administered at 14 weeks of age) and measles (administered at nine months of age) for children aged 12–35 months from the DHS. We defined a child as having received DTP3 if they had received three doses and measles if they had received the first dose. This could be recorded by a mark on a vaccine card or a mother’s response. DTP3 and measles vaccine coverage were found to be highly correlated at the country level (Figure S2) and therefore we used DTP3 vaccination status as an indicator of access to vaccination.

All DHS and MIS collect information on ITNs, therefore data on ITN access and usage could be linked to data on vaccination uptake at the individual level. We defined a child as using an ITN if they slept in the house on the previous night and used either an ITN, or both an ITN and untreated net, that previous night. We defined a child as having access to a net if they slept in the house on the previous night and had a mosquito bed net for sleeping. We did not consider IRS due to the low global percentage of households protected by this method.

Data on *P. falciparum* malaria prevalence and the proportion of children seeking treatment for fever were not consistently available in all DHS surveys and therefore could only be matched at the country or admin-1 level, using MIS data where possible (Table S1). Treatment coverage was defined as the proportion of children aged between 12–59 months for whom fever was reported within the previous two weeks and who sought medical treatment. *P. falciparum* malaria prevalence was based on rapid diagnostic test (RDT) results in children aged between 12–59 months. The prevalence data were not included in the survey data sets for two countries (Cameroon and Zambia), therefore these values were obtained from the corresponding reports (Table S1). All country and admin-1 coverage estimates were adjusted using the reported sample weights according to the DHS guidance [19].

We extracted data on gender, age, rural/urban status, the highest level of the mother’s education and the wealth index to explore additional determinants underlying variation in access to vaccines and malaria interventions. These variables are included in the DHS and were therefore matched to vaccination status, ITN access and ITN usage at the individual level. All data were extracted using the *rdhs* package in R software [22].

### Analysis

We defined six groups based on the vaccination status, ITN access and ITN usage for each child:

1. Did not receive the DTP3 vaccine and did not have access to or sleep under an ITN;
2. Did not receive the DTP3 vaccine and had access to, but did not sleep under, an ITN;
3. Did not receive the DTP3 vaccine but did sleep under an ITN;
4. Received the DTP3 vaccine but did not have access to or sleep under an ITN;
5. Received the DTP3 vaccine had access to, but did not sleep under, an ITN;
6. Received the DTP3 vaccine and slept under an ITN.

The first group we term “missed children” since these children do not have access to an ITN and are also unlikely to benefit from the introduction of the malaria vaccine introduced via the EPI. The fourth and fifth groups we term “prospective children” since these children are not currently using ITNs but could be accessed via EPI to receive the malaria vaccine. Differences in the distribution of children by group between countries were evaluated using a Chi-squared test.

We fitted a multinomial mixed effect model for nominal (unordered) outcomes to explore the determinants of overlap in vaccine coverage and ITN access and usage. This allowed groups 2–6 to be contrasted against the reference group (the “missed children”), while accounting for variation at the country level using a random intercept [23, 24]. The following variables were considered during the model building process: age of the child; malaria prevalence of the region in which the child lived; sex of the child; mother’s education level; and whether the region in which a child lived was classified as rural or urban. We did not consider clustering at the survey cluster level because there were very few observations associated with each survey cluster. Prevalence was not found to be a significant variable and was subsequently removed from the model. These analyses were undertaken using SAS version 9.4 [25].

We estimated the number of children aged 12–59 months at risk of malaria for each country by multiplying the proportion of children in each group at the at the admin-1 level by the at-risk population for each admin-1 unit and aggregating it to the country level. Demographic and population data were obtained from the United Nations World Population Prospects 2015 and 2016 projections respectively [26]. The population-at-risk was then calculated using at-risk proportions obtained by masking regions designated outside the spatial limits of *P. falciparum* [27]. We then estimated the number of cases that could be averted by the vaccine in prospective children using the at-risk populations of the prospective group who received the DTP3 vaccine but did not use an ITN (groups 4 and 5). We also calculated the number of cases averted for those who already have a vaccine and use an ITN (group 6). We assumed 100% coverage of the malaria vaccine across the children who received DTP3 in these three groups and that the vaccine reduces the clinical incidence of malaria across this age group by 39% irrespective of whether they used an ITN [28]. We calculated the number of cases averted at the admin-1 level and aggregated at the country level. To obtain the clinical incidence prior to vaccine introduction, we used a model-based relationship between parasite prevalence and clinical incidence (Figure S2) to translate the estimates of parasite prevalence by RDT in children younger than five years from the corresponding DHS or MIS [29].

## Results

Figure 1 shows the country-level relationship between DTP3 vaccine coverage and three malaria intervention coverage indicators (ITN usage, ITN access and the proportion of fever cases seeking treatment). Across the majority of countries, vaccination coverage was higher than any of the selected malaria intervention coverage indicators. The highest levels of malaria intervention coverage were access to ITNs; with eight of the 20 of the countries analysed having higher ITN access compared to vaccination coverage (Angola, Benin, Cote d’Ivoire, DRC, Guinea, Mali, Nigeria and Uganda). However, this difference was only substantial (>10%) in three of these countries (Benin, Malawi and Nigeria). Furthermore, there was a consistent gap between ITN access and ITN usage in all countries. ITN usage was therefore lower than vaccination coverage in all but three countries (Benin, DRC and Mali). Only one country (Angola) had lower vaccine coverage than proportion of children seeking treatment for fever.

**Figure 1:**
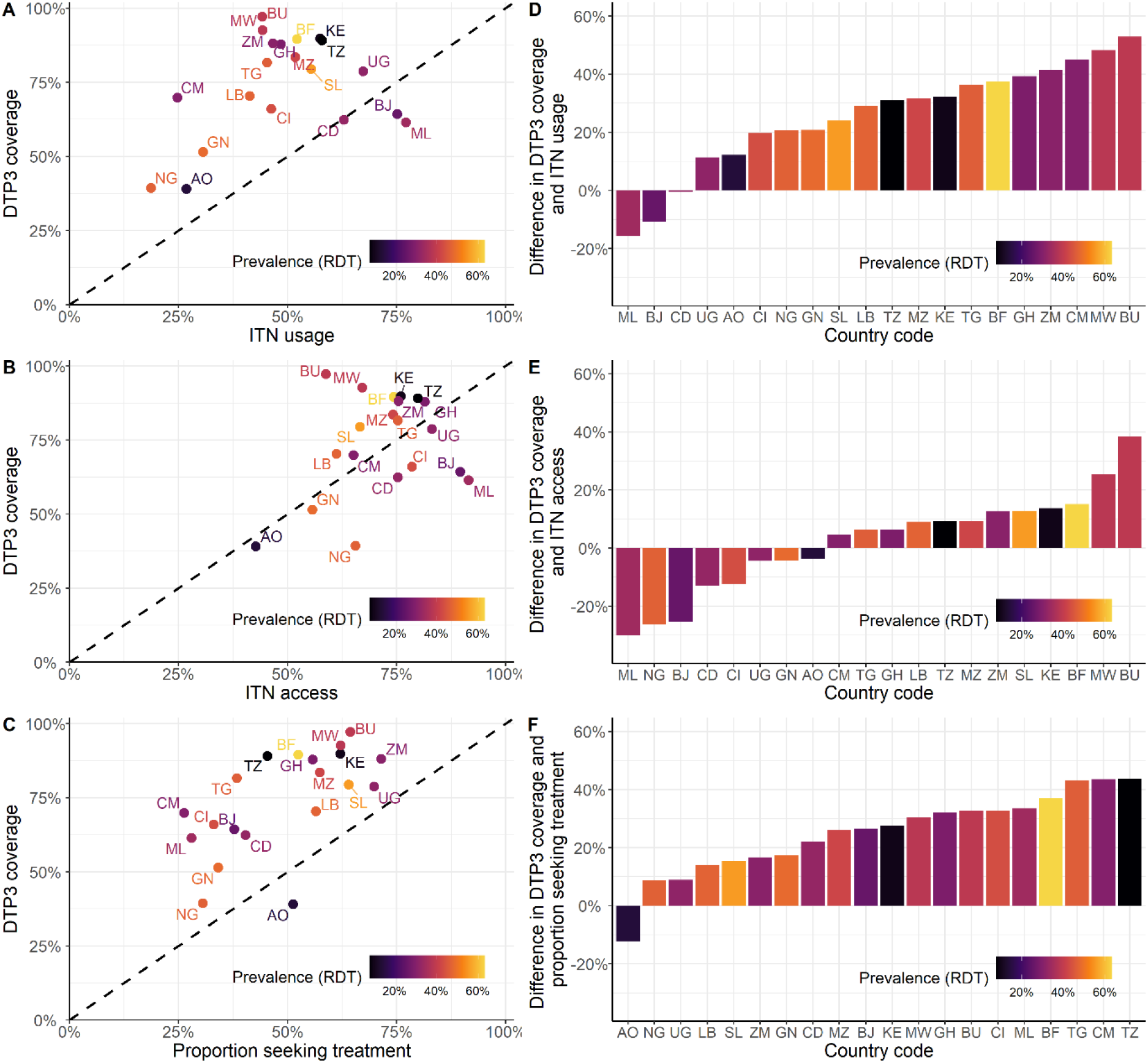
Comparison of ITN usage, ITN access, proportion of children seeking treatment and DTP3 coverage. The colour gradient represents *P. falciparum* prevalence in children aged 12–59 months. The countries shown are: AO – Angola, BF – Burkina Faso, BJ – Benin, BU – Burundi, CD - Democratic Republic of Congo, CI – Cote d’Ivoire, CM – Cameroon, GH – Ghana, GN – Guinea, KE – Kenya, LB – Liberia, ML – Mali, MW – Malawi, MZ – Mozambique, NG – Nigeria, SL – Sierra Leone, TG – Togo, TZ – Tanzania, UG – Uganda and ZM – Zambia. (A) (B) and (C) show ITN usage, ITN access and the proportion of children seeking treatment versus DTP3 coverage respectively. D) Difference in DTP3 coverage and ITN usage ranked in order of increasing difference with the bars coloured by *P. falciparum* prevalence. E) Difference in DTP3 coverage and ITN access ranked in order of increasing difference with the bars coloured by *P. falciparum* prevalence. F) Difference in DTP3 coverage and proportion of children seeking treatment with the bars coloured by *P. falciparum* prevalence.

Overall, 34% of children did not use ITNs but received the DTP3 vaccine, and 35% of children both used ITNs and received the DTP3 vaccine (Table 1). There was significant variation in the overlap between ITN use and DTP3 vaccine uptake between countries (chi-squared test, p<0.001). The highest proportion of children who received no DTP3 vaccine and did not use an ITN were in south-western Africa, whereas the lowest proportion were in the south east of Africa (Figure 2A). In contrast, the highest proportion of children who received the DTP3 vaccine and did not use an ITN were in south-eastern Africa and the lowest were south-western Africa (Figure 2B). Examining unvaccinated children, 16% of children used ITNs but were not vaccinated, and 15% of children did not receive either intervention.

**Table 1:**
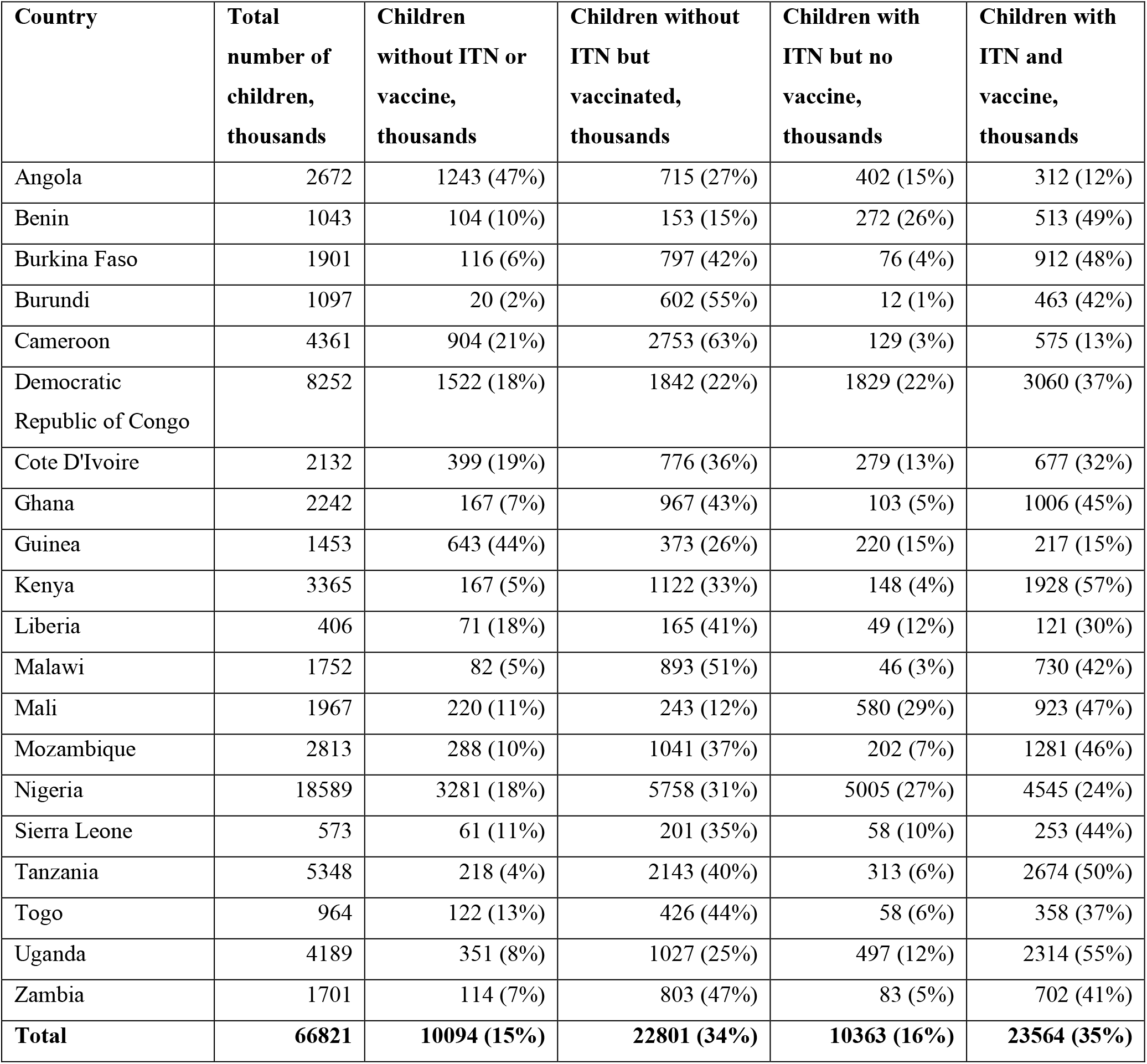
Numbers of children aged 12–59 months at risk of malaria and receiving different intervention combinations (DTP3 vaccination and/or an ITN). The percentage is relative to the total for that country. The analysis was performed at the admin-1 unit level and the total numbers of children were then aggregated at the country level.

**Figure 2:**
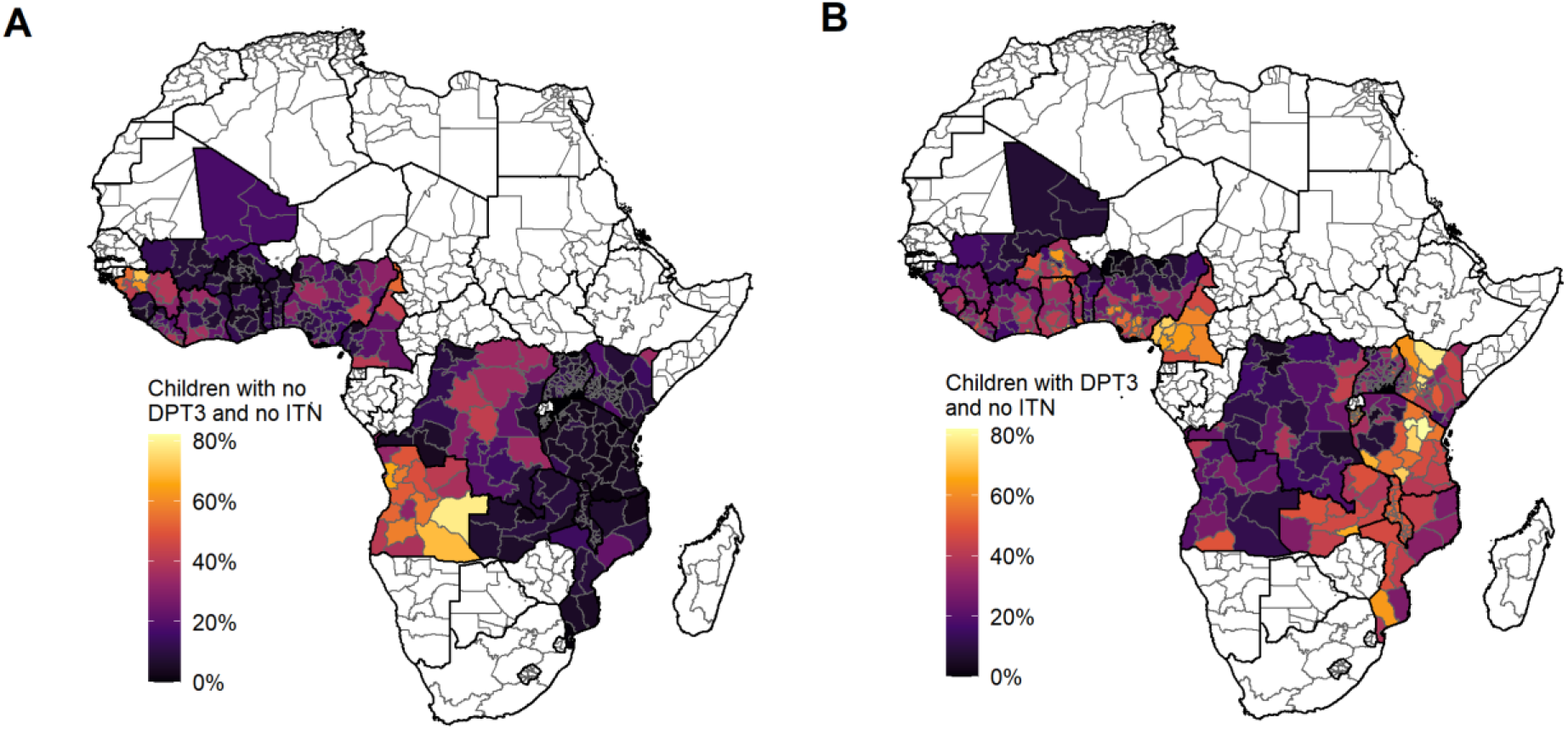
Proportion of the population aged 12–59 months in each country who have do not use an ITN (includes both access and does not use, and no access) and have various vaccination statuses. A) Proportion of children who have not been vaccinated with the DTP3 vaccine and did not use an ITN. B) Proportion of children who have been vaccinated with the DTP3 vaccine and did not use an ITN. The countries shown are Angola, Burkina Faso, Benin, Burundi, Democratic Republic of Congo, Cote d’Ivoire, Cameroon, Ghana, Guinea, Kenya, Liberia, Mali, Malawi, Mozambique, Nigeria, Sierra Leone, Togo, Tanzania, Uganda and Zambia.

We estimated that in the 20 countries analysed there are 33 million children at risk of malaria who currently do not use an ITN (Table 1). Of these, 23 million (70%) are in the “prospective” group that did receive the DTP3 vaccine and hence could potentially receive protection from the malaria vaccine under existing distribution channels. Cameroon, Democratic Republic of Congo, Kenya, Mozambique, Nigeria, Tanzania and Uganda each have more than one million children in this group. However, Angola, Democratic Republic of Congo and Nigeria each have more than one million children in the “missed children” category who did not receive the DTP3 vaccine and do not have access to a net. Across the 20 countries, approximately the same number of children in the “prospective” group, who do not currently use an ITN, had access to an ITN than did not despite individual countries having very different levels of overall access to ITNs. This pattern was repeated in the “missed children” group.

By vaccinating all children in the “prospective” group (children who receive DTP3 but are not protected by an ITN), an estimated 9.7 million malaria cases could be averted each year across these 20 countries (Table 2). An additional 10.8 million cases could be averted by vaccinating those 23 million children who both use an ITN and have the DTP3 vaccine. Despite the relatively low vaccination coverage levels in Nigeria, 30% of total cases averted would be averted by vaccinating children in this country with a further 22% in two additional countries – Cameroon and the Democratic Republic of Congo.

**Table 2:** Estimated malaria cases averted. The numbers of cases, and percentage relative to the total across all countries, that could be averted annually in 12–59-month-old children if universal coverage of the RTS,S/AS01 vaccine was achieved, for those who received the DPT3 vaccine in each country. These estimated are calculated for individual admin-1 units and aggregated at the country level.

| Country | Total cases averted in thousands | Cases averted in children using an ITN in thousands (percentage of total averted across all countries) | Cases averted in children without an ITN in thousands (percentage of total averted across all countries) |
| --- | --- | --- | --- |
| Angola | 162 | 54 (0%) | 109 (1%) |
| Benin | 282 | 220 (1%) | 63 (0%) |
| Burkina Faso | 1326 | 729 (4%) | 597 (3%) |
| Burundi | 583 | 237 (1%) | 346 (2%) |
| Cameroon | 1359 | 219 (1%) | 1140 (6%) |
| Democratic Republic of Congo | 2307 | 1420 (7%) | 888 (4%) |
| Cote D'Ivoire | 834 | 433 (2%) | 400 (2%) |
| Ghana | 836 | 465 (2%) | 371 (2%) |
| Guinea | 396 | 160 (1%) | 236 (1%) |
| Kenya | 536 | 399 (2%) | 137 (1%) |
| Liberia | 222 | 94 (0%) | 128 (1%) |
| Malawi | 930 | 416 (2%) | 514 (3%) |
| Mali | 575 | 465 (2%) | 110 (1%) |
| Mozambique | 1251 | 692 (3%) | 559 (3%) |
| Nigeria | 5684 | 2887 (14%) | 2797 (14%) |
| Sierra Leone | 351 | 204 (1%) | 147 (1%) |
| Tanzania | 522 | 338 (2%) | 184 (1%) |
| Togo | 578 | 264 (1%) | 314 (2%) |
| Uganda | 1317 | 889 (4%) | 428 (2%) |
| Zambia | 491 | 245 (1%) | 246 (1%) |
| **Total** | **20542** | **10827 (53%)** | **9715 (47%)** |

The association between demographic and socioeconomic variables and the different intervention coverage groups are shown in Figure 3. Older children (aged 24–35 months) were less likely to have received the DTP3 vaccine and use an ITN compared to younger children (aged 12–23 months). There were notable differences in intervention coverage between rural and urban populations; those that had access to or used a net were more likely to reside in rural areas compared to the missed children regardless of vaccination status (p=0.034) whereas those that received the DTP3 vaccine, but did not have access to a net, were more likely to reside in urban areas compared to the missed children (p=0.006). The mother’s education status was strongly associated with intervention uptake with membership of all intervention groups that either used an ITN and/or received the DTP3 vaccine (groups 3, 4, 5 and 6) being significantly associated with higher levels of education (p<0.001). Furthermore, membership of the group that did not receive the vaccine and had access to a net but did not use it (group 2) was significantly less likely in those that had received higher education compared to those that received little or no education. The wealth index was also a strong predictor of intervention coverage. Membership of all intervention groups that either had received the DTP3 vaccine, or owned and/or used a net, was significantly associated with higher wealth levels after we controlled for the urban/rural divide. Those receiving higher incomes were approximately twice as likely to access both nets and vaccines (groups 5 and 6) compared to those in the lower wealth quintile (p<0.001). The numbers of children in each group are provided in Table S2.

**Figure 3:**
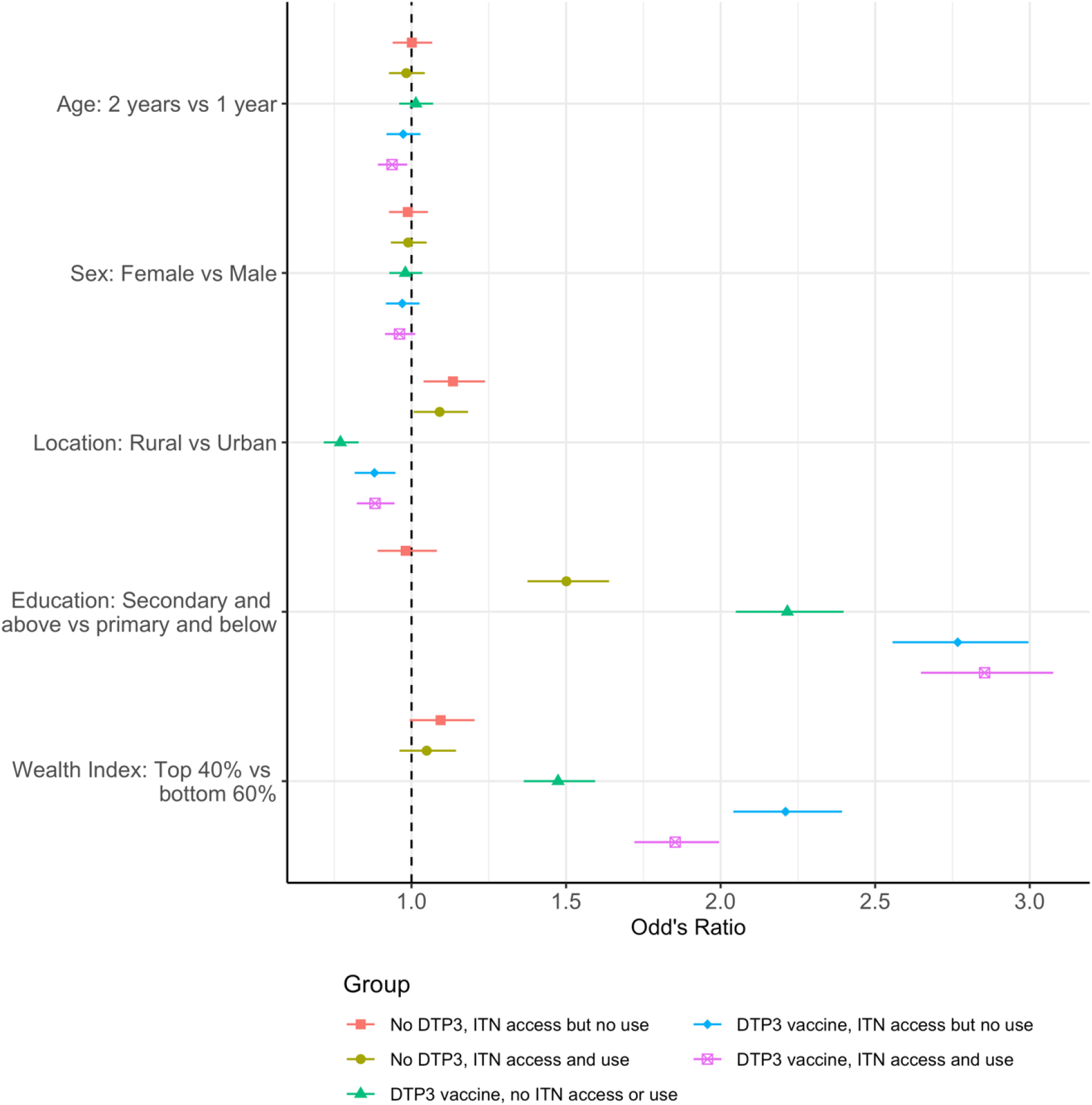
The relationship between demographic and socioeconomic variables and intervention coverage. Odd’s ratio estimates, 95% confidence levels and p values are shown for each of the predictors. All ratios are comparative to group 1 where children did not receive the DTP3 vaccine and have no access to an ITN (the “missed children”). Children in group 2 did not receive the DTP3 vaccine and had access to, but did not sleep under, an ITN; children in group 3 did not receive the DTP3 vaccine but did sleep under an ITN; children in group 4 received the DTP3 vaccine but did not have access to or sleep under an ITN; children in group 5 received the DTP3 vaccine and had access to, but did not sleep under, an ITN; and children in group 6 received the DTP3 vaccine and slept under an ITN.

## Discussion

The substantial declines in the burden of malaria in sub-Saharan Africa since 2010 have been attributed primarily to the rapid increase in access and usage of ITNs alongside slower but significant improvements in access to first-line treatment [30]. However, in more recent years the coverage of both interventions has plateaued, with the most recent household surveys analysed here demonstrating sub-optimal levels of ITN usage in many countries (at or below 50%) and even lower rates of treatment seeking for fever. In contrast, supported by the establishment of Gavi, The Vaccine Alliance in 2000 and the World Health Organization’s EPI, uptake of childhood vaccination has steadily increased, although the past decade has seen some stagnation [14, 31]. As demonstrated by our analysis, vaccine coverage is high in most of the 20 countries studied here, with only two countries (Angola and Guinea) reporting under 50% uptake in their most recent DHS. Furthermore, our results demonstrate that coverage of vaccines administered via the EPI is substantially higher than usage of ITNs or fever treatment-seeking rates in the majority of countries, which is consistent with other research [7]. This creates an opportunity to consider roll-out strategies for the introduction of a malaria vaccine that are distinct to implementation programs for other malaria interventions, to maximise impact by building on the wider reach of the EPI programme.

As noted elsewhere, the utilisation of malaria interventions and uptake of vaccination were found to be strongly associated with demographic and socioeconomic indicators [5–8, 12, 13]. Perhaps not surprisingly given the differing distribution mechanisms, those accessing and using ITNs were more likely to reside in rural areas whereas as higher uptake of the DTP3 vaccine was associated with urban areas, possibly as a result of better access to healthcare facilities. Uptake of vaccination was also strongly associated with both the mother’s educational level and the wealth quintile, although as noted elsewhere, even at lower levels of education and wealth, the coverage of vaccination remained high [13]. Access to ITNs and ITN usage were also both strongly associated with the mother’s education status and wealth; however, having access to a net but not using it whilst also not being vaccinated with DTP3 was more strongly associated with lower educational status of mothers than with wealth.

We estimate that there are currently 33 million children in these 20 countries who are not using an ITN. Of these, 23 million (70% of the total children without an ITN) are estimated to have received the DTP3 vaccine and hence could be reached by the EPI programme. If the RTS,S malaria vaccine were made available to just these 23 million children, we estimate that 9.7 million cases (or 0.44 cases per vaccinated child) could be averted each year (assuming all children who receive the DTP3 vaccine also receive the RTS,S). If the vaccine was also administered to those children with an ITN and who were vaccinated (24 million additional children), we estimate an additional 10.8 million cases or (0.47 cases per vaccinated child) could also be averted. Around 40% of the total cases would be averted in the Democratic Republic of Congo and Nigeria, countries which are currently contributing large proportions of malaria cases worldwide. This alone could represent a substantial reduction in the global malaria burden, reducing *P. falciparum* malaria cases by approximately 4%. However, there are still 9.8 million children across our countries of interest who would remain unprotected by either intervention; these “missed children” should remain the focus of initiatives to improve equity in access to both malaria interventions and vaccination.

There are several limitations to this analysis. First, we were only able to undertake the analysis for 20 of the 27 malaria-endemic countries of interest within sub-Saharan Africa. Those countries for which data were not available contributed around 10% of the Africa malaria burden in 2018 and therefore also remain an important target for both malaria interventions and vaccination [2]. Second, due to the different survey designs, we were not able to link vaccination status and access to treatment at the individual level. Given that both rely on access to health services, it is likely that these may be correlated, although levels of access to treatment remain well below vaccination rates. An alternative equity dimension that may be relevant to consider when targeting malaria interventions could be the potential for access to rapid treatment since both severe disease incidence and malaria mortality have been associated with the time taken to reach care [32, 33]. Third, to estimate the impact of the RTS,S vaccine on malaria burden we used the mean estimate of vaccine efficacy across the phase 3 trial sites over a four-year period (39%). As usage of ITNs during the trial was very high, this likely represents an under-estimate of the true impact of the vaccine in this group of prospective children. A study using RTS,S trial and bed net usage in Malawi estimated that vaccinating a child in urban Lilongwe without a bed net could prevent 1.09 malaria cases, versus 0.67 for a child with a bed net. In rural Lilongwe, 2.59 and 1.59 malaria cases could be averted, respectively [34]. Furthermore, in taking this simple approach to estimating vaccine impact, we have ignored the differences in vaccine efficacy by endemicity that were observed in the trial, and the potential age-shifting of cases that would likely occur as a result of reduced exposure to infection [35]. Fourth, we also assume universal malaria vaccine coverage for children who receive the DTP3 vaccine, which results in our estimate of cases averted being an upper bound. The RTS,S vaccine is delivered as a four-dose schedule, with the first and third doses aligning with existing EPI contact points. Therefore, particularly in the early phase of vaccine introduction, it is possible that vaccine coverage could be lower than that of DTP3. Projected vaccine coverage will be informed by data from the pilot studies as they progress. Finally, our analysis is based on self-reported vaccination status and ITN use from household surveys across different years; as such they are not directly comparable with estimates produced by WHO (for malaria interventions and vaccination) and UNICEF (for vaccination) which are obtained by triangulating data from a number of sources.

## Conclusions

In summary, broadly high levels of childhood vaccination across malaria-endemic countries in sub-Saharan Africa, including in high *P. falciparum* prevalence regions where take-up of current interventions remains sub-optimal, provide an opportunity to maximise the impact of childhood malaria vaccination. Older children were less likely to have received the DTP3 vaccine and use an ITN compared to younger children and the higher the levels of mother’s education and wealth indexes, the greater the intervention coverage. Our study also highlights the considerable number of children who are not currently accessing routine childhood immunisations or core malaria interventions and who should be the focus of health equity initiatives in order to improve access. These combined findings allow for the identification of populations that could benefit most from the introduction of a childhood malaria vaccine and could be used to devise strategies for future malaria vaccine implementation.

## Data Availability

The data that support the findings of this study are available from the DHS Program repository (https://dhsprogram.com/) and can be accessed upon registration and application.

## List of abbreviations

WHO: World Health Organization
GTS: World Health Organization Global Technical Strategy for Malaria
ITN: insecticide-treated net
DTP3: diphtheria-tetanus-pertussis vaccine dose 3
IRS: indoor residual spraying
RTS,S: RTS,S/AS01 malaria vaccine for children
DHS: Demographic and Health Surveys
MIS: Malaria Indicator Surveys
EPI: Expanded Programme on Immunization
MAP: Malaria Atlas Project
*Pf*PR_2-10_: *P. falciparum* parasite prevalence in 2–10-year-old individuals
IQR: inter-quartile range.

## Authors’ contributions

ABH and ACG conceived and designed the study. HJTU, ABH and LM extracted the data and ran the analyses. All authors interpreted the findings. HJTU and ABH wrote the first draft of the paper. All authors contributed to and approved the final version of the manuscript.

