## Supplementary Information for "Analysis of the potential for a malaria vaccine to reduce gaps in malaria intervention coverage"

**Table S1: Countries eligible for inclusion in the analysis and surveys from which data were extracted.** All surveys are available for download from the Demographic and Health Surveys website [1].

| **Country** | **Surveys used** |
|---|---|
| Angola (AO) | 2015 DHS |
| Benin (BJ) | 2012 DHS |
| Burkina Faso (BF) | 2010 DHS, 2014 MIS |
| Burundi (BU) | 2016 DHS |
| Cameroon (CM) | 2011 DHS |
| Cote d'Ivoire (CI) | 2012 DHS |
| Democratic Republic of Congo (CD) | 2013 DHS |
| Ghana (GH) | 2014 DHS, 2016 MIS |
| Guinea (GN) | 2012 DHS |
| Kenya (KE) | 2014 DHS, 2015 MIS |
| Liberia (LB) | 2013 DHS, 2016 MIS |
| Malawi (MW) | 2015 DHS, 2017 MIS |
| Mali (ML) | 2012 DHS, 2015 MIS |
| Mozambique (MZ) | 2015 AIS |
| Nigeria (NG) | 2013 DHS, 2015 MIS |
| Sierra Leone (SL) | 2013 DHS, 2016 MIS |
| Tanzania (TZ) | 2015 DHS, 2017 MIS |
| Togo (TG) | 2013 DHS, 2017 MIS |
| Uganda (UG) | 2016 DHS |
| Zambia (ZM) | 2013 DHS, 2015 MIS |

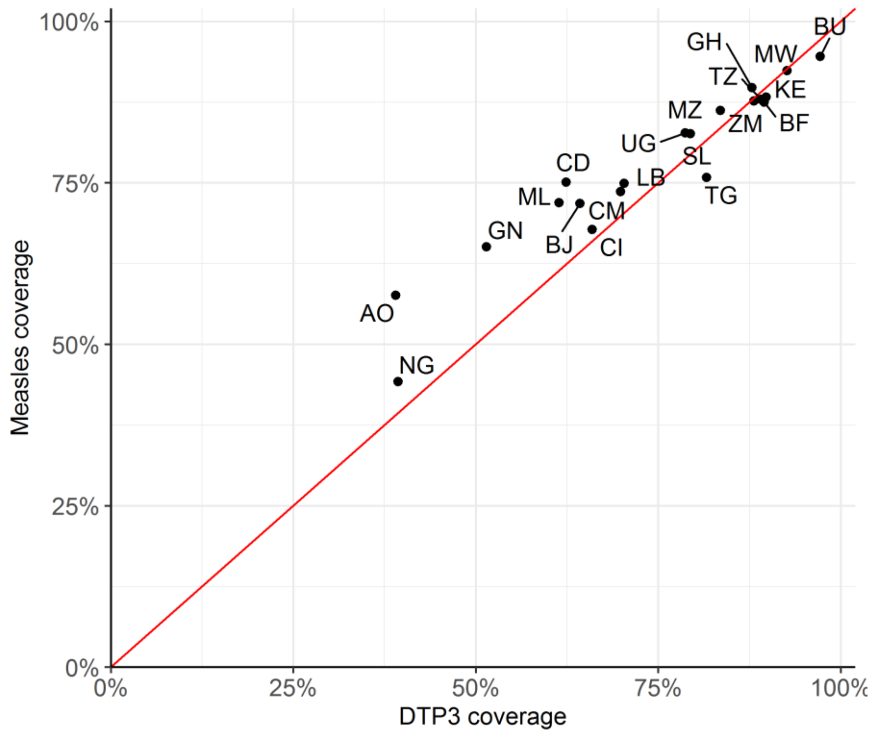


**Figure S1: Coverage of the DTP3 vaccine at the country level, compared to measles vaccine coverage.** Spearman's correlation coefficient: 0.950. The red line represents correlation of 1.0. Countries shown: AO – Angola, BF – Burkina Faso, BJ – Benin, BU – Burundi, CD - Democratic Republic of Congo, CI – Cote d'Ivoire, CM – Cameroon, GH – Ghana, GN – Guinea, KE – Kenya, LB – Liberia, ML – Mali, MW – Malawi, MZ – Mozambique, NG – Nigeria, SL – Sierra Leone, TG – Togo, TZ – Tanzania, UG - Uganda and ZM – Zambia.

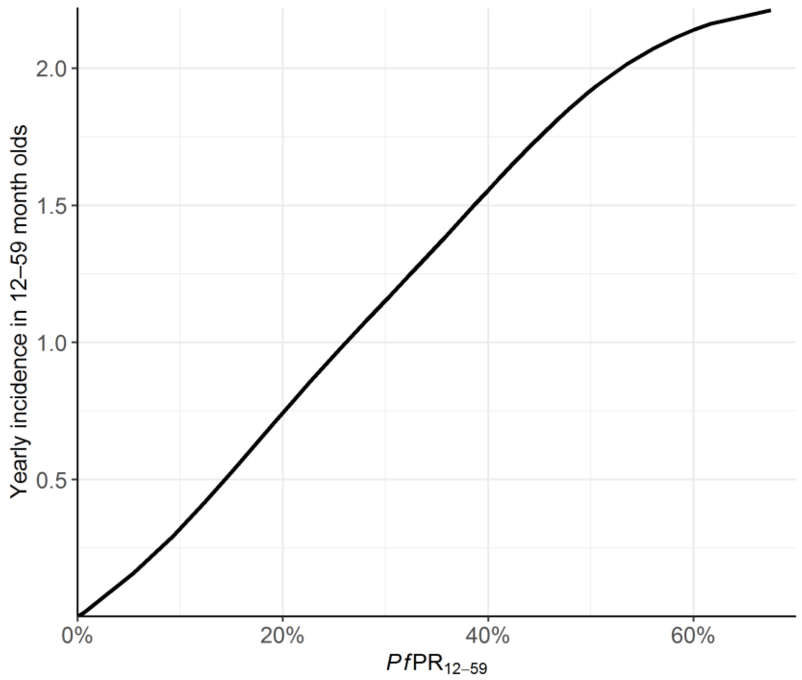


**Figure S2: Yearly clinical incidence of malaria as a function of the *P. falciparum* parasite prevalence in children aged 12–59 months (*Pf*PR12–59).** Clinical incidence is the yearly number of events per child aged 12–59 months. This relationship was computed using a deterministic mathematical model for malaria transmission [2].

**Table S2: Numbers of children and proportions of multinomial regression predictors in each country.**

| Country code | Number of children (thousands) | Sex | | Age | | Urban/ Rural | | Mother's education | | Wealth Index | |
|---|---|---|---|---|---|---|---|---|---|---|---|
| | | Male | Female | One | Two | Urban | Rural | None & Primary | Secondary & Tertiary | Bottom 60% | Top 40% |
| Angola | 5451 | 50% | 50% | 52% | 48% | 56% | 44% | 71% | 29% | 78% | 23% |
| Burkina Faso | 4817 | 51% | 49% | 52% | 48% | 37% | 63% | 89% | 12% | 64% | 36% |
| Benin | 5461 | 51% | 49% | 51% | 49% | 24% | 76% | 94% | 6% | 61% | 39% |
| Burundi | 4979 | 50% | 50% | 52% | 48% | 16% | 84% | 87% | 13% | 60% | 39% |
| Democratic Republic of Congo | 2293 | 49% | 51% | 53% | 47% | 42% | 58% | 65% | 35% | 64% | 36% |
| Cote d'Ivoire | 2814 | 50% | 50% | 50% | 50% | 34% | 66% | 89% | 10% | 70% | 30% |
| Cameroon | 6831 | 50% | 50% | 50% | 50% | 30% | 70% | 65% | 35% | 69% | 31% |
| Ghana | 2496 | 52% | 48% | 52% | 48% | 31% | 69% | 89% | 11% | 63% | 37% |
| Guinea | 2255 | 52% | 48% | 50% | 50% | 41% | 59% | 54% | 46% | 71% | 28% |
| Kenya | 8055 | 51% | 49% | 50% | 50% | 32% | 68% | 73% | 27% | 71% | 29% |
| Liberia | 2699 | 52% | 48% | 53% | 47% | 32% | 68% | 79% | 20% | 83% | 17% |
| Mali | 6492 | 49% | 51% | 50% | 50% | 16% | 84% | 79% | 22% | 65% | 36% |
| Malawi | 3608 | 50% | 50% | 51% | 49% | 26% | 74% | 90% | 10% | 58% | 41% |
| Mozambique | 1977 | 48% | 52% | 51% | 49% | 36% | 64% | 79% | 21% | 56% | 44% |
| Nigeria | 11154 | 51% | 49% | 52% | 48% | 34% | 66% | 65% | 35% | 64% | 36% |
| Sierra Leone | 4012 | 49% | 51% | 51% | 49% | 30% | 70% | 81% | 19% | 64% | 36% |
| Togo | 4032 | 50% | 50% | 53% | 47% | 24% | 76% | 81% | 19% | 62% | 37% |
| Tanzania | 2676 | 50% | 50% | 52% | 48% | 30% | 70% | 78% | 22% | 68% | 32% |
| Uganda | 5814 | 51% | 49% | 50% | 50% | 19% | 81% | 75% | 25% | 66% | 33% |
| Zambia | 5092 | 50% | 50% | 51% | 49% | 37% | 63% | 66% | 34% | 71% | 29% |
